# An Evaluation of DMR-Informed Fine-Tuning of Tissue Array–Pretrained CpGPT for Gastrointestinal Cancer Classification Using cfDNA Targeted Methylation

**DOI:** 10.64898/2026.07.26.26358979

**Authors:** Yili Xu, Qianxi Feng

## Abstract

**Background:** Cell-free DNA (cfDNA) methylation profiling is promising for minimally invasive cancer detection, but its translation is limited by high-dimensional data, modest cfDNA cohort sizes, and the difficulty of defining biologically grounded feature sets. CpGPT, a transformer-based DNA methylation foundation model pretrained on large-scale tissue methylation array datasets, may enable transfer of learned methylation representations to data-limited cfDNA applications. We evaluated a differentially methylated region (DMR)-informed framework for fine-tuning CpGPT for gastrointestinal cancer classification using plasma cfDNA methylation data.

**Methods:** Plasma cfDNA methylation data were obtained from the EpiPanGI Dx cohort, including 254 gastrointestinal cancer samples and 46 non-cancer controls. Tumor and matched normal tissue methylation data included 967 tumor-normal pairs across 18 cancer types from The Cancer Genome Atlas (TCGA). Tissue-derived and cfDNA-derived DMRs were identified independently and intersected to define a candidate set of 20,499 CpG sites. CpGPT was evaluated in a technical sensitivity analysis using the previously published 896-CpG panGI panel and in DMR-informed fine-tuning using the 20,499-CpG candidate set. Performance was compared with radial-kernel support vector machine (SVM radial), elastic net logistic regression, gradient boosting machine (GBM), and Random Forest models using identical data partitions.

**Results:** In the technical sensitivity analysis using the previously published panGI panel, the CpGPT base configuration achieved a mean test area under the receiver operating characteristic curve (AUROC) of 0.9799 (95% CI, 0.9654–0.9944) across four fixed random seeds. Individual test AUROCs ranged from 0.9597 to 0.9927, while neighboring configurations achieved mean test AUROCs of 0.9661–0.9716. Using the DMR-informed candidate set, CpGPT achieved a mean test AUROC of 0.9904 (95% CI, 0.9715–1.0000) across three split-runs. Mean test AUROCs were 0.9573 for SVM radial, 0.9420 for elastic net, 0.8460 for GBM, and 0.8408 for Random Forest. Cancer type-specific mean test AUROCs ranged from 0.9600 for pancreatic adenocarcinoma to 1.0000 for esophageal squamous cell carcinoma, colorectal cancer, and esophageal adenocarcinoma.

**Conclusions:** DMR-informed CpGPT fine-tuning achieved high internal discrimination and a higher mean test AUROC than the evaluated classical machine learning models. Integrating tissue and plasma cfDNA methylation evidence provides a biologically constrained feature space for adapting a tissue array-pretrained foundation model to cfDNA classification. Independent external validation, clinically representative control populations, and further feature-set reduction are needed before translation into a targeted cfDNA assay.

## Introduction

Cell-free DNA (cfDNA) has emerged as a promising minimally invasive analyte for cancer detection and characterization. Among the molecular features measurable in cfDNA, DNA methylation is particularly informative because methylation patterns are highly cell-type specific and can reflect both the tissue source of circulating DNA and cancer-associated epigenetic alterations. Methylation-based analyses have demonstrated the potential to detect cancer-associated signals and classify their tissue of origin across multiple malignancies ^[1,2]^.

These properties are especially relevant to gastrointestinal cancers, several of which lack effective blood-based screening approaches for average-risk populations. The EpiPanGI Dx study used tissue methylation discovery followed by targeted bisulfite sequencing of plasma cfDNA to develop cancer-specific, pan-gastrointestinal, and tissue-of-origin methylation classifiers^[3]^. The study demonstrated the potential of plasma DNA methylation for detecting hepatocellular carcinoma, pancreatic adenocarcinoma, esophageal adenocarcinoma, gastric cancer, esophageal squamous cell carcinoma, and colorectal cancer. The corresponding publicly available cfDNA dataset also provides a resource for evaluating alternative computational approaches to gastrointestinal cancer classification.

Despite this promise, developing robust predictive models from cfDNA methylation profiles remains challenging. Methylation datasets may contain hundreds of thousands of CpG sites, whereas cfDNA studies frequently include comparatively modest sample sizes. The low fraction of circulating tumor DNA in plasma, together with variation in sequencing coverage, further complicates the identification of reproducible cancer-associated signals. Models trained on high-dimensional, minimally filtered CpG matrices may therefore be susceptible to overfitting and may be difficult to interpret or convert into practical targeted assays. Conventional machine learning methods, including regularized regression, support vector machines, random forests, and gradient boosting models, provide important benchmarks but generally operate on predefined CpG feature vectors without explicitly integrating genomic position and local sequence context.

Biologically informed feature construction offers one strategy for addressing these limitations. Tissue-derived differentially methylated regions (DMRs) identify loci associated with epigenetic remodeling in tumors, whereas cfDNA-derived DMRs identify methylation differences that remain detectable in circulation. Integrating these complementary sources of evidence can reduce the initial feature space to loci supported by both tumor biology and plasma cfDNA measurements. This approach extends the tissue-to-plasma biomarker discovery strategy used in EpiPanGI Dx while providing a defined candidate CpG space for downstream foundation-model fine-tuning and machine learning comparisons^[3]^.

Transformer-based foundation models offer an additional strategy for modeling high-dimensional methylation data. By pretraining on large collections of methylation profiles, these models can learn reusable representations that incorporate relationships among methylation state, genomic position, and sequence context. CpGPT was developed as a foundation model for DNA methylation analysis and supports methylation reconstruction, embedding generation, and fine-tuning for downstream classification tasks^[4]^. MethylGPT independently demonstrated that transformer-based pretraining can capture local and higher-order methylation patterns and support diverse phenotype-classification applications^[5]^. These studies suggest that methylation foundation models may provide advantages over models trained de novo in datasets with limited sample sizes.

In this study, we developed and evaluated a DMR-informed CpGPT fine-tuning framework for cfDNA-based gastrointestinal cancer classification. We hypothesized that intersecting tumor tissue-derived and plasma cfDNA-derived DMRs would define a biologically constrained candidate CpG feature space that could support efficient adaptation of a pretrained methylation foundation model. We first evaluated the technical sensitivity of CpGPT fine-tuning using the previously developed 896-CpG panGI methylation panel. We then fine-tuned CpGPT using a broader set of 20,499 DMR-informed candidate CpG sites and compared its performance with radial-kernel support vector machine (SVM radial), elastic net logistic regression, gradient boosting machine (GBM), and Random Forest models using identical data partitions. Finally, we evaluated CpGPT performance across six gastrointestinal cancer types. This study provides a proof-of-concept evaluation of DMR-informed foundation-model adaptation for cfDNA methylation-based cancer classification.

## Methods

### Study Design and Overview

This retrospective computational study evaluated a differentially methylated region (DMR)-informed framework for adapting a pretrained DNA methylation foundation model to cfDNA-based gastrointestinal cancer classification. All analyses used previously generated, publicly available methylation data; no new participants were recruited, and no new biospecimens were collected.

The analytical workflow comprised three components. First, tissue-derived and plasma cfDNA-derived DMRs were integrated to construct a biologically constrained candidate CpG feature set. Second, CpGPT was fine-tuned for binary cancer-versus-control classification using either the previously developed pan-gastrointestinal (pan-GI) cancer methylation panel or the broader DMR-informed candidate set. Third, CpGPT performance was compared with four classical machine learning models using identical feature sets and data partitions.

The primary performance metric was the area under the receiver operating characteristic curve (AUROC). Secondary metrics included area under the precision-recall curve (AUPRC), stability across random seeds and model configurations, and cancer type-specific classification performance.

### Data Sources and Study Cohorts

Plasma cfDNA methylation data were obtained from the National Center for Biotechnology Information Gene Expression Omnibus (NCBI GEO) under accession number GSE149438. The dataset was generated as part of the EpiPanGI Dx study, which used targeted bisulfite sequencing to profile plasma cfDNA from patients with gastrointestinal cancers and non-cancer controls^[3]^.

The cfDNA cohort consisted of 300 plasma samples, including 254 cancer samples and 46 non-cancer control samples. The cancer samples comprised hepatocellular carcinoma (HCC), pancreatic adenocarcinoma (PDAC), esophageal adenocarcinoma (EAC), gastric cancer (GC), esophageal squamous cell carcinoma (ESCC), and colorectal cancer (CRC). Cancer type-specific sample counts are presented in **Table 1**.

**Table 1.**
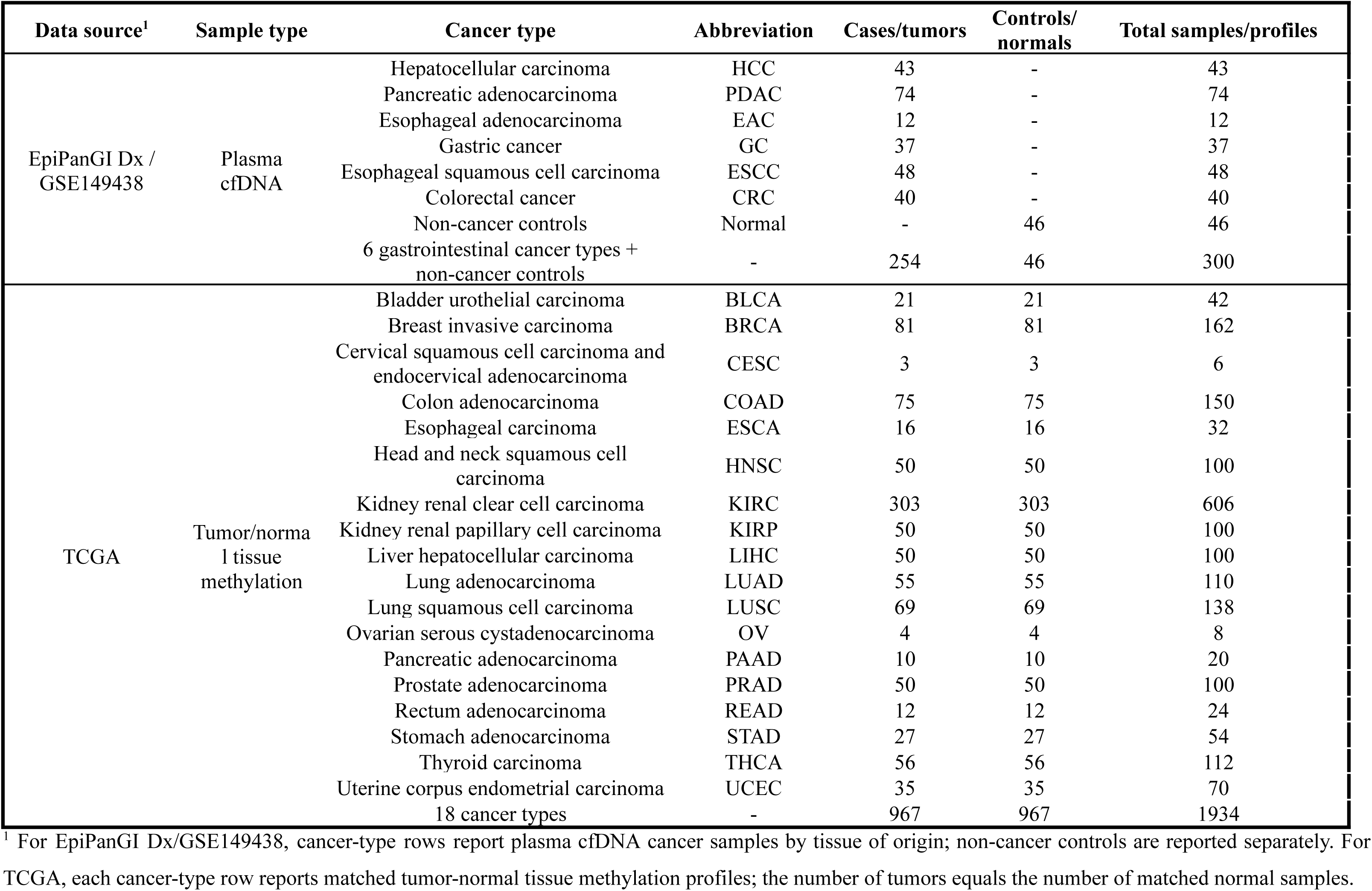
Cohort and feature-set overview by cancer type.

| Data source<sup>1</sup> | Sample type | Cancer type | Abbreviation | Cases/tumors | Controls/<br>normals | Total samples/profiles |
| --- | --- | --- | --- | --- | --- | --- |
| EpiPanGI Dx /<br>GSE149438 | Plasma<br>cfDNA | Hepatocellular carcinoma | HCC | 43 | - | 43 |
|  |  | Pancreatic adenocarcinoma | PDAC | 74 | - | 74 |
|  |  | Esophageal adenocarcinoma | EAC | 12 | - | 12 |
|  |  | Gastric cancer | GC | 37 | - | 37 |
|  |  | Esophageal squamous cell carcinoma | ESCC | 48 | - | 48 |
|  |  | Colorectal cancer | CRC | 40 | - | 40 |
|  |  | Non-cancer controls | Normal | - | 46 | 46 |
|  |  | 6 gastrointestinal cancer types +<br>non-cancer controls | - | 254 | 46 | 300 |
| TCGA | Tumor/normal<br>tissue<br>methylation | Bladder urothelial carcinoma | BLCA | 21 | 21 | 42 |
|  |  | Breast invasive carcinoma | BRCA | 81 | 81 | 162 |
|  |  | Cervical squamous cell carcinoma and<br>endocervical adenocarcinoma | CESC | 3 | 3 | 6 |
|  |  | Colon adenocarcinoma | COAD | 75 | 75 | 150 |
|  |  | Esophageal carcinoma | ESCA | 16 | 16 | 32 |
|  |  | Head and neck squamous cell<br>carcinoma | HNSC | 50 | 50 | 100 |
|  |  | Kidney renal clear cell carcinoma | KIRC | 303 | 303 | 606 |
|  |  | Kidney renal papillary cell carcinoma | KIRP | 50 | 50 | 100 |
|  |  | Liver hepatocellular carcinoma | LIHC | 50 | 50 | 100 |
|  |  | Lung adenocarcinoma | LUAD | 55 | 55 | 110 |
|  |  | Lung squamous cell carcinoma | LUSC | 69 | 69 | 138 |
|  |  | Ovarian serous cystadenocarcinoma | OV | 4 | 4 | 8 |
|  |  | Pancreatic adenocarcinoma | PAAD | 10 | 10 | 20 |
|  |  | Prostate adenocarcinoma | PRAD | 50 | 50 | 100 |
|  |  | Rectum adenocarcinoma | READ | 12 | 12 | 24 |
|  |  | Stomach adenocarcinoma | STAD | 27 | 27 | 54 |
|  |  | Thyroid carcinoma | THCA | 56 | 56 | 112 |
|  |  | Uterine corpus endometrial carcinoma | UCEC | 35 | 35 | 70 |
|  |  | 18 cancer types | - | 967 | 967 | 1934 |

<sup>1</sup> For EpiPanGI Dx/GSE149438, cancer-type rows report plasma cfDNA cancer samples by tissue of origin; non-cancer controls are reported separately. For TCGA, each cancer-type row reports matched tumor-normal tissue methylation profiles; the number of tumors equals the number of matched normal samples.

For tissue-based methylation discovery, tumor and matched normal tissue methylation data were obtained from The Cancer Genome Atlas (TCGA)^[6]^. The tissue dataset included 967 tumor-normal pairs, corresponding to 1,934 tissue methylation profiles across 18 cancer types. The cancer types and their respective sample counts are summarized in **Table 1**.

### cfDNA Methylation Data Processing

Plasma cfDNA methylation profiles were generated in the source study using targeted bisulfite sequencing. After adapter and low-quality base trimming, sequencing reads were aligned to the hg19 human reference genome using BSMAP version 2.90^[7]^. CpG-level methylation ratios were calculated using methratio.py from the BSMAP package.

Methylation levels were represented as beta values, defined as the number of methylated reads divided by the total number of reads covering each CpG site. CpG measurements supported by fewer than four sequencing reads were excluded. CpG sites with greater than 20% missingness across samples were removed from the present analysis.

For model development, missing methylation values were imputed using the median beta value for each CpG site. Imputation values were estimated using the training partition only and subsequently applied to the corresponding validation and test partitions to prevent information leakage.

### DMR Discovery and Candidate CpG Construction

Tissue-derived and cfDNA-derived DMRs were identified independently before genomic integration.

### Tissue-Derived DMR Discovery

Within each TCGA cancer type, methylation profiles from tumor tissues were compared with those from matched normal tissues. Differential methylation analysis was performed using the limma framework^[8]^. Candidate DMRs were required to contain at least three CpG sites, have an absolute mean methylation difference greater than 0.10, and have a Benjamini-Hochberg false discovery rate-adjusted P-value less than 0.05^[9]^.

To prioritize methylation changes observed across tumor types, tissue-derived DMRs were retained if they were identified in at least two cancer types. CpG sites contained within the retained tissue-derived DMR intervals were then enumerated, resulting in 344,566 tissue-derived CpG sites for subsequent integration.

### cfDNA-Derived DMR Discovery

Plasma cfDNA methylation profiles from cancer cases were compared with those from non-cancer controls. Region-level analysis was used to identify coordinated methylation differences across adjacent CpG sites.

Candidate cfDNA-derived DMRs were required to contain at least three CpG sites, have an absolute mean methylation difference greater than 0.10, and have a Benjamini-Hochberg-adjusted P-value less than 0.05. Additional filters required a minimum region length of 150 base pairs, a consistent direction of methylation difference among CpG sites within each region, and sufficient sequencing coverage for reliable methylation estimation.

CpG sites contained within the qualifying cfDNA-derived DMR intervals were enumerated, resulting in 23,277 cfDNA-derived CpG sites for genomic integration.

### Integration of Tissue- and cfDNA-Derived DMRs

Tissue-derived and cfDNA-derived DMR intervals were intersected using BEDTools^[10]^. Genomic loci supported by both tumor-normal tissue methylation differences and cancer-versus-control plasma cfDNA methylation differences were considered high-confidence cancer-associated methylation loci.

CpG sites located within overlapping tissue and cfDNA DMR intervals were retained. This procedure generated a DMR-informed candidate feature set of 20,499 CpG sites, which was used for CpGPT fine-tuning and classical machine learning benchmarking.

### CpGPT Foundation Model

CpGPT is a transformer-based foundation model pretrained on large collections of DNA methylation profiles^[4]^. The model incorporates methylation-state, genomic-position, and local DNA-sequence information to learn contextual representations of CpG methylation patterns. It supports methylation reconstruction, embedding generation, and supervised fine-tuning for downstream classification tasks.

Fine-tuning in the present study was initialized from the pretrained Cancer-100M CpGPT checkpoint. We used the large CpGPT architecture, comprising 512-dimensional sequence and methylation embeddings, a hidden dimension of 512, 32 transformer layers, and 16 attention heads. For each CpG site, a 2,001-bp local DNA-sequence context was encoded. The resulting CpG representations were processed by the Transformer++ backbone and a task-specific condition decoder. Binary cancer-versus-control classification was optimized using binary cross-entropy loss.

### CpGPT Fine-Tuning Experiments

#### Technical Sensitivity Analysis Using the Previously Published panGI Feature Panel

The purpose of this analysis was to evaluate the sensitivity of CpGPT fine-tuning to modest changes in optimization settings when applied to a fixed, previously published 896-CpG pan-gastrointestinal methylation panel. This analysis was designed as a technical sensitivity analysis rather than an independent validation of the feature panel or an estimate of clinical generalizability.

All 300 cfDNA samples were evaluated through repeated internal data partitions. Four neighboring fine-tuning configurations were evaluated. The base configuration used a learning rate of 8×10^−6^, a condition-loss weight of 3.0, dropout of 0.02, an early-stopping patience of 18 validation evaluations, and a maximum of 3,000 training steps. Three one-parameter variants were assessed relative to the base configuration: dropout03 increased dropout to 0.03, lr6e6 reduced the learning rate to 6×10^−6^, and cond4 increased the condition-loss weight to 4.0. Each configuration was evaluated using four prespecified random seeds: 2601, 2602, 2603, and 2604. Internal test-set AUROC was summarized across seeds to assess the sensitivity of model performance to stochastic variation and modest hyperparameter perturbations.

#### DMR-Informed CpGPT Fine-Tuning

Cancer-versus-control labels from the cfDNA cohort were intentionally used during candidate-feature discovery because the objective of this stage was supervised biomarker prioritization for future targeted-assay development, rather than unsupervised representation learning. Specifically, cfDNA-derived DMR discovery was conducted using all 300 samples in the source cohort, comprising 254 cancer cases and 46 non-cancer controls, and the resulting cfDNA DMRs were intersected with the tissue-derived DMRs to define the 20,499-CpG candidate set. The resulting 20,499-CpG set should therefore be interpreted as an assay-development candidate feature set derived from the full source cohort. Performance estimates obtained through subsequent internal data partitioning may remain influenced by cohort-level feature discovery and should not be interpreted as fully independent validation.

CpGPT was fine-tuned using the pretrained Cancer-100M checkpoint. The DMR-informed configuration used a learning rate of 8×10^−6^, a condition-loss weight of 3.0, dropout of 0.02, weight decay of 0.1, a batch size of 2, and a maximum of 4,000 training steps. The data loader allowed a maximum input length of 10,000 CpG sites per sample; when more than 10,000 eligible sites were available, a subset of the 20,499 candidate CpGs was used for each model input. Three independent train-validation-test partitions were generated using split seeds 42, 123, and 456. The same split assignments were retained for CpGPT and all classical machine learning models to permit direct performance comparison. Model selection and training decisions were based only on the training and validation partitions, and final performance was evaluated in the held-out test partition.

#### Classical Machine Learning Benchmarks

Four classical machine learning models were evaluated: radial-kernel support vector machine (SVM radial), elastic net logistic regression, gradient boosting machine (GBM), and Random Forest^[11–14]^. Models were implemented in R using the caret framework and associated model-specific packages^[15]^.

Each model used the same 20,499-CpG candidate feature set and the same training, validation, and test partitions as CpGPT. All preprocessing parameters were estimated within the training partition. Missing beta values were imputed using training-set medians, CpG sites with zero or near-zero variance were removed, and CpG beta values were centered and scaled for models requiring standardized inputs.

For elastic net logistic regression, the mixing parameter and regularization strength were selected through validation-set performance or cross-validation within the training partition. For SVM radial models, the cost and radial-kernel width parameters were tuned. Random Forest tuning considered the number of variables sampled at each split and terminal-node size. GBM tuning considered the number of trees, interaction depth, learning rate, and minimum number of observations per terminal node.

#### Cancer Type-Specific Performance

Cancer type-specific CpGPT performance was evaluated for hepatocellular carcinoma (HCC), pancreatic adenocarcinoma (PDAC), esophageal adenocarcinoma (EAC), gastric cancer (GC), esophageal squamous cell carcinoma (ESCC), and colorectal cancer (CRC).

Within each split-run, predicted probabilities from test samples of a given cancer type were compared with predicted probabilities from non-cancer controls in the corresponding test partition. AUROC and AUPRC were calculated separately for each cancer type. Cancer type-specific results were then summarized across the three split-runs.

### Model Evaluation and Statistical Analysis

AUROC was the primary model-performance metric. AUPRC was evaluated as a secondary threshold-independent metric. Where calculated, threshold-dependent metrics included accuracy, sensitivity, specificity, precision, recall, and F1 score.

For each model or cancer type, performance was summarized as the arithmetic mean across independent runs. Descriptive 95% confidence intervals (CIs) across runs were calculated as: 95% CI = mean ± 1.96 × SD/√n, where SD was the standard deviation across runs and n was the number of runs. For the panGI technical sensitivity analysis, n = 4; for DMR-informed fine-tuning, classical machine learning comparisons, and cancer type-specific analyses, n = 3. Confidence limits were bounded to the valid metric range of 0 to 1. These intervals quantify variability across random-seed or data-split runs and do not represent participant-level bootstrap confidence intervals.

### Reproducibility and Code Availability

All analyses were conducted using R and Python. CpGPT fine-tuning was performed using the CpGPT codebase and pretrained Cancer-100M checkpoint. Classical machine learning benchmarks were implemented in R using caret and associated model-specific packages.

Prespecified random seeds were used for data partitioning and model training. Training-derived preprocessing parameters were applied unchanged to validation and test data. Code used for methylation preprocessing, DMR integration, CpGPT fine-tuning, classical machine learning benchmarking, cancer type-specific evaluation, and performance summarization will be made publicly available at publication or upon reasonable request, subject to the data-use requirements of the source datasets.

## Results

### Study Cohorts and DMR-Informed Candidate CpG Construction

The analysis included 300 plasma cfDNA methylation samples from the EpiPanGI Dx cohort, comprising 254 gastrointestinal cancer samples and 46 non-cancer controls. Cancer samples included 74 pancreatic adenocarcinomas, 48 esophageal squamous cell carcinomas, 43 hepatocellular carcinomas, 40 colorectal cancers, 37 gastric cancers, and 12 esophageal adenocarcinomas. The tissue methylation dataset included 1,934 profiles from 967 matched tumor-normal pairs across 18 TCGA cancer types (**Table 1**).

Application of the prespecified tissue DMR criteria identified qualifying DMR intervals containing 344,566 CpG sites. Independent analysis of plasma cfDNA identified qualifying cancer-versus-control DMR intervals containing 23,277 CpG sites. Genomic intersection of the tissue-derived and cfDNA-derived DMR intervals yielded 20,499 CpG sites supported by both tumor tissue methylation differences and circulating cfDNA methylation differences. These CpG sites constituted the DMR-informed candidate feature set used for CpGPT fine-tuning and classical machine learning benchmarking (**Figure 1**).

**Figure 1.**
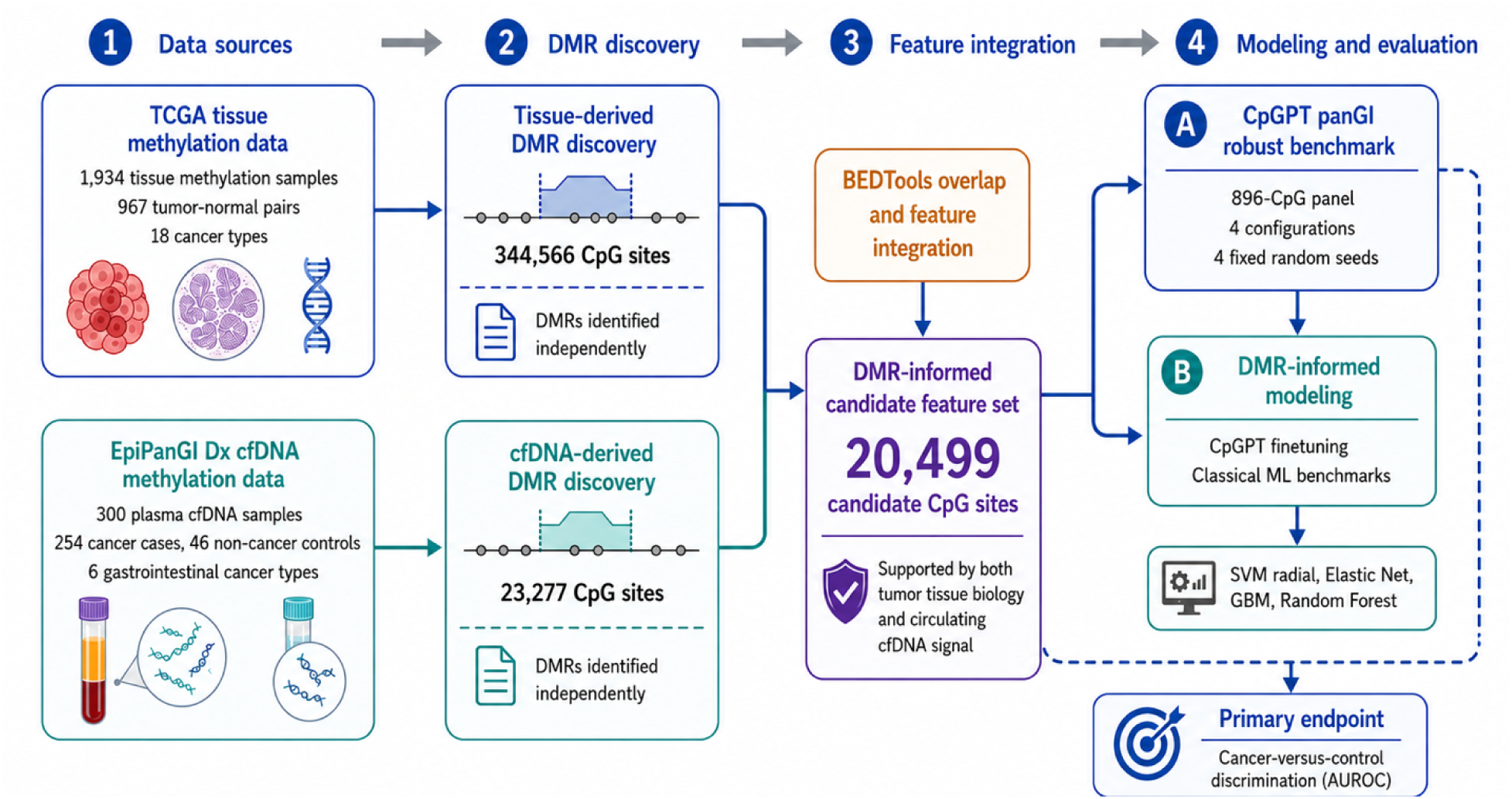
Overview of the DMR-informed CpGPT framework for cfDNA cancer classification. The study workflow integrates tumor tissue methylation data and plasma cfDNA methylation data to construct a biologically informed CpG feature space for cancer-versus-control classification. Tissue-derived differentially methylated regions (DMRs) were identified from TCGA tumor-normal methylation data, while cfDNA-derived DMRs were identified from the EpiPanGI Dx plasma cfDNA dataset. Genomic interval overlap was used to integrate tissue and cfDNA DMR evidence, producing a DMR-informed candidate feature set of 20,499 CpG sites. CpGPT was evaluated in two modeling settings: a technical sensitivity analysis using the previously published 896-CpG panGI panel across multiple configurations and random seeds, and DMR-informed modeling using the 20,499-CpG candidate set. Classical machine learning benchmarks included radial-kernel support vector machine (SVM radial), elastic net logistic regression, gradient boosting machine (GBM), and Random Forest. The primary endpoint was cancer-versus-control discrimination, measured by the area under the receiver operating characteristic curve (AUROC).

### CpGPT Performance Was Consistent Across Settings in the Technical Sensitivity Analysis

CpGPT was first evaluated using the 896-CpG panGI methylation panel across four neighboring hyperparameter configurations and four fixed random seeds per configuration (**Table 2** and **Figure 2A**).

**Figure 2.**
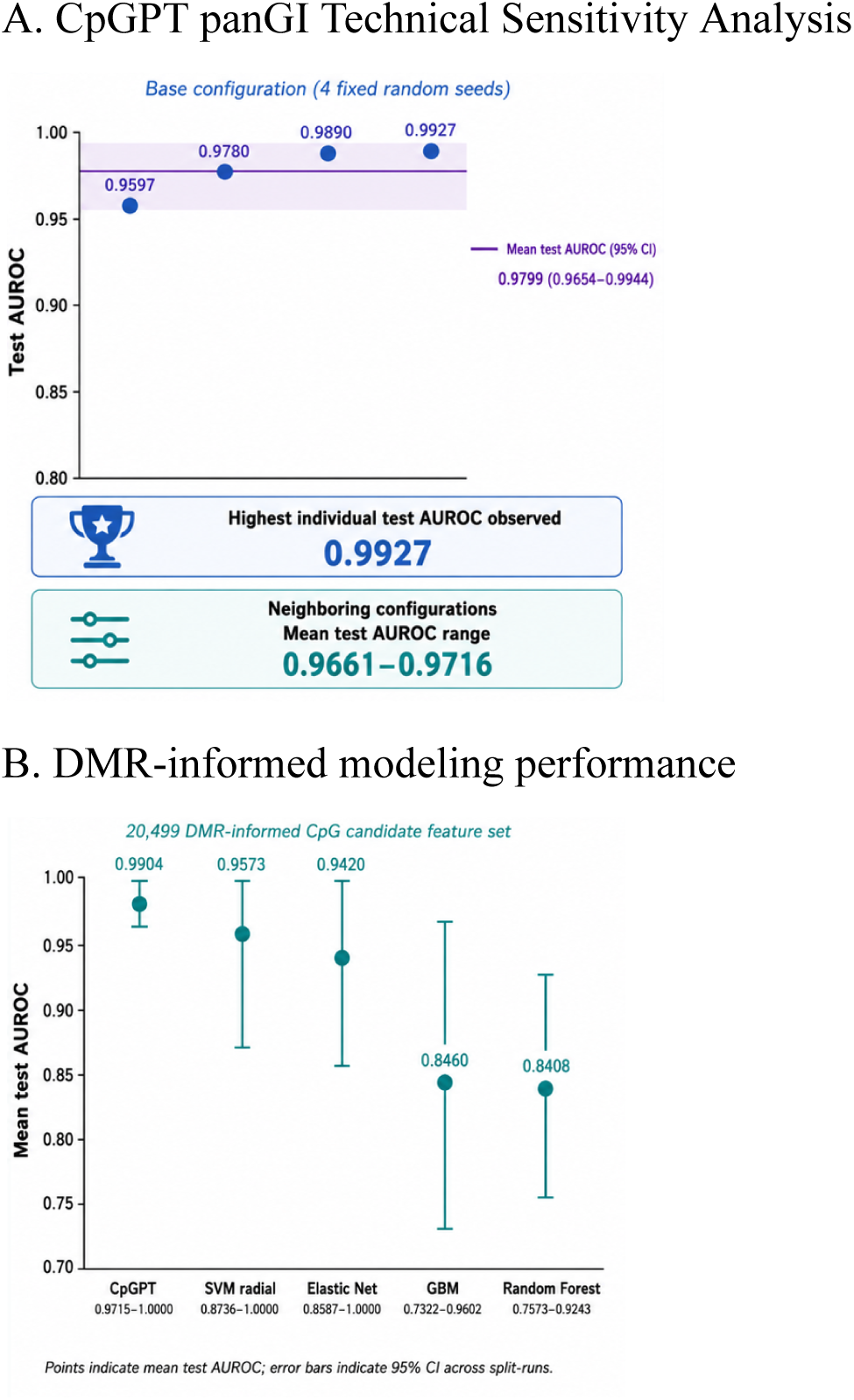
CpGPT and classical machine learning model performance. **(A)** CpGPT performance in the technical sensitivity analysis using the previously published panGI panel using the 896-CpG panGI methylation panel. Under the base configuration, CpGPT was evaluated across four fixed random seeds, yielding individual test AUROCs of 0.9597, 0.9780, 0.9890, and 0.9927. The mean test AUROC was 0.9799 (95% CI 0.9654–0.9944). The highest individual test AUROC observed was 0.9927, and neighboring configurations achieved mean test AUROCs ranging from 0.9661 to 0.9716. **(B)** modeling performance using the 20,499 DMR-informed CpG candidate feature set. CpGPT achieved the highest mean test AUROC across three split-runs (0.9904, 95% CI 0.9715–1.0000), followed by radial-kernel support vector machine (SVM radial, 0.9573, 95% CI 0.8736–1.0000), elastic net logistic regression (0.942, 95% CI 0.8587–1.0000), gradient boosting machine (GBM, 0.846, 95% CI 0.7322–0.9602), and Random Forest (0.8408, 95% CI 0.7573–0.9243). Points indicate mean test AUROC, and error bars indicate 95% CI across split-runs.

**Table 2.** CpGPT technical sensitivity analysis using the previously published panGI feature panel.

| Configuration | Learning rate | Condition loss weight | Dropout | Mean test AUROC (95% CI)<sup>1</sup> | Individual test AUROCs |
| --- | --- | --- | --- | --- | --- |
| Base | $8 \times 10^{-6}$ | 3 | 0.02 | 0.9799 (0.9654–0.9944) | 0.9597, 0.9780, 0.9890, 0.9927 |
| dropout03 | $8 \times 10^{-6}$ | 3 | 0.03 | 0.9661 (0.9451–0.9872) | 0.9487, 0.9744, 0.9487, 0.9927 |
| lr6e6 | $6 \times 10^{-6}$ | 3 | 0.02 | 0.9716 (0.9565-0.9867) | 0.9560, 0.9707, 0.9670, 0.9927 |
| cond4 | $8 \times 10^{-6}$ | 4 | 0.02 | 0.9689 (0.9493-0.9885) | 0.9634, 0.9744, 0.9451, 0.9927 |

<sup>1</sup> Mean and 95% confidence interval (CI) are summarized across four fixed random seeds where available.

Under the base configuration, individual test AUROCs were 0.9597, 0.9780, 0.9890, and 0.9927. The mean test AUROC was 0.9799 (95% CI, 0.9654–0.9944). The dropout03, lr6e6, and cond4 configurations achieved mean test AUROCs of 0.9661 (95% CI, 0.9451–0.9872), 0.9716 (95% CI, 0.9565–0.9867), and 0.9689 (95% CI, 0.9493–0.9885), respectively. The highest individual test AUROC observed across the benchmark configurations was 0.9927.

Within this internal technical sensitivity analysis, CpGPT produced consistently high discrimination across the evaluated random seeds and modest changes in learning rate, dropout, and condition-loss weight. These findings describe the sensitivity of fine-tuning behavior within the source cohort and do not constitute independent validation of the previously published panel.

### DMR-Informed CpGPT Fine-Tuning Achieved High Held-Out Performance

CpGPT was next fine-tuned using the 20,499 DMR-informed candidate CpG sites across three independent data splits. Validation AUROCs were 0.8942, 1.0000, and 0.9520, corresponding to a mean validation AUROC of 0.9487 (95% CI, 0.8887–1.0000).

Test AUROCs were 1.0000, 1.0000, and 0.9712 across the three split-runs. The resulting mean test AUROC was 0.9904 (95% CI, 0.9715–1.0000), and the mean test AUPRC was 0.9960 (95% CI, 0.9950–0.9970) (**Table 3**).

**Table 3.** Performance comparison between CpGPT and classical machine learning baselines.

| Model | Validation AUROC<sup>1</sup> | Test AUROC<sup>1</sup> | Test AUPRC<sup>1</sup> |
| --- | --- | --- | --- |
| CpGPT | 0.9487<br>(0.8887–1.0000) | 0.9904<br>(0.9715–1.0000) | 0.9960<br>(0.9950–0.9970) |
| SVM radial | 0.9367<br>(0.8126–1.0000) | 0.9573<br>(0.8736–1.0000) | 0.9827<br>(0.9488–1.0000) |
| Elastic Net | 0.9300<br>(0.7928–1.0000) | 0.9420<br>(0.8587–1.0000) | 0.9805<br>(0.9494–1.0000) |
| GBM | 0.8767<br>(0.7917–0.9617) | 0.8460<br>(0.7322–0.9602) | 0.9550<br>(0.9122–0.9978) |
| Random Forest | 0.7933<br>(0.5946–0.9920) | 0.8408<br>(0.7573–0.9243) | 0.9475<br>(0.8985–0.9965) |

<sup>1</sup> Mean and 95% confidence interval (CI) are summarized across three split-runs. The 95% CIs were bounded to the valid metric range of 0 to 1.

### CpGPT Achieved the Highest Mean Test AUROC Among the Evaluated Models

Using identical DMR-informed CpG features and data partitions, CpGPT was compared with SVM radial, elastic net logistic regression, GBM, and Random Forest models (**Table 3** and **Figure 2B**).

CpGPT achieved the highest mean test AUROC at 0.9904 (95% CI, 0.9715–1.0000). Among the classical machine learning models, SVM radial achieved the highest mean test AUROC at 0.9573 (95% CI, 0.8736–1.0000), followed by elastic net at 0.9420 (95% CI, 0.8587–1.0000). GBM and Random Forest achieved mean test AUROCs of 0.8460 (95% CI, 0.7322–0.9602) and 0.8408 (95% CI, 0.7573–0.9243), respectively.

Mean test AUPRC was 0.9960 (95% CI, 0.9950–0.9970) for CpGPT, 0.9827 (95% CI, 0.9488–1.0000) for SVM radial, 0.9805 (95% CI, 0.9494–1.0000) for elastic net, 0.9550 (95% CI, 0.9122–0.9978) for GBM, and 0.9475 (95% CI, 0.8985–0.9965) for Random Forest. These comparisons were descriptive across the three split-runs.

### CpGPT Performance Was High Across Gastrointestinal Cancer Types

Cancer type-specific performance was evaluated by comparing test samples from each gastrointestinal cancer type with the non-cancer controls in the corresponding test partition (**Table 4** and **Figure 3**).

**Figure 3.**
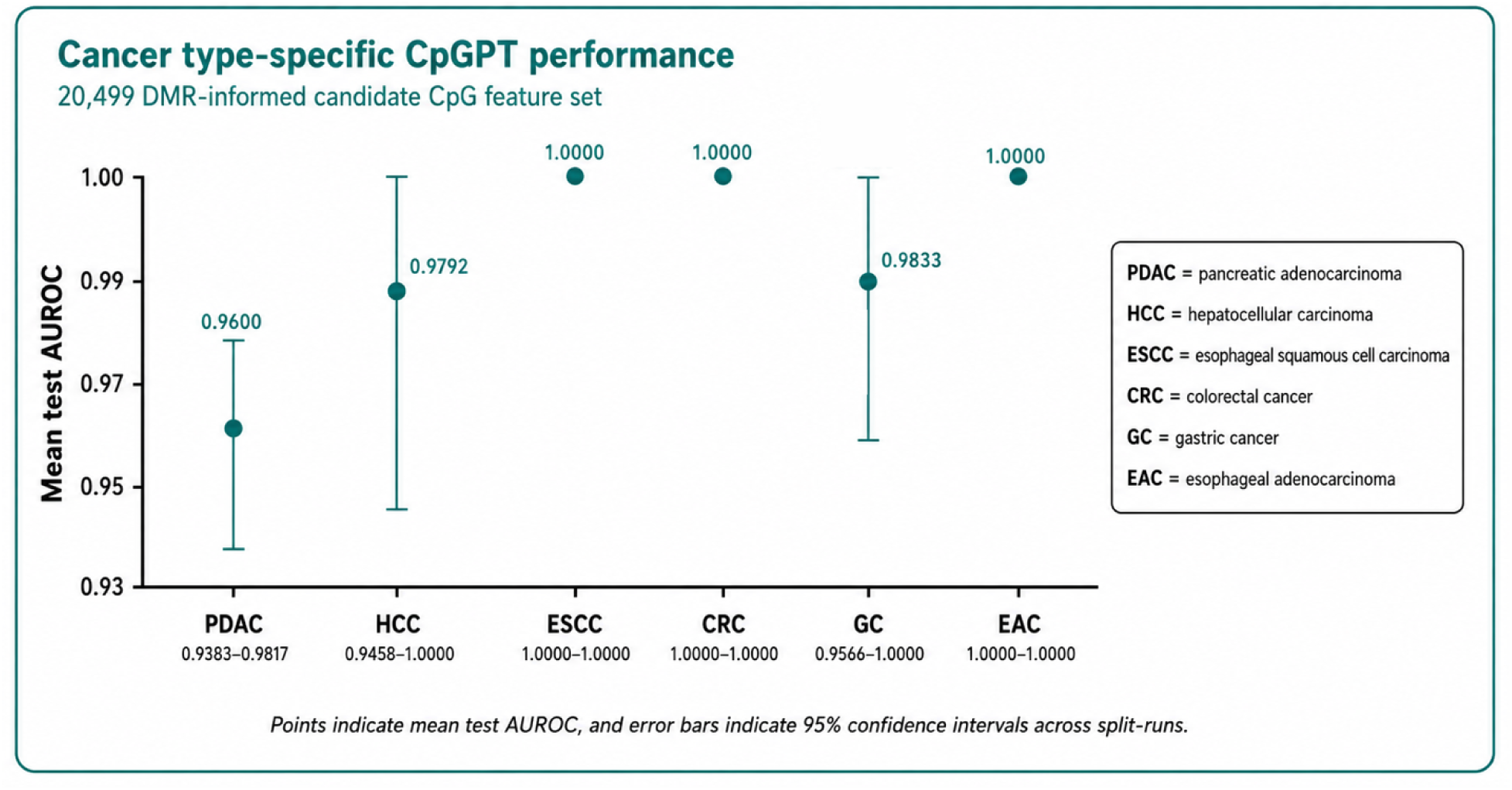
CpGPT performance by gastrointestinal cancer type. Cancer type-specific CpGPT performance was evaluated using the 20,499 DMR-informed candidate CpG feature set across three split-runs. Mean test AUROC was 0.9600 for pancreatic adenocarcinoma (PDAC; 95% CI, 0.9383–0.9817), 0.9792 for hepatocellular carcinoma (HCC; 95% CI, 0.9458–1.0000), 1.0000 for esophageal squamous cell carcinoma (ESCC; 95% CI, 1.0000–1.0000), 1.0000 for colorectal cancer (CRC; 95% CI, 1.0000–1.0000), 0.9833 for gastric cancer (GC; 95% CI, 0.9566–1.0000), and 1.0000 for esophageal adenocarcinoma (EAC; 95% CI, 1.0000–1.0000). Points indicate mean test AUROC, and error bars indicate 95% confidence intervals across split-runs.

**Table 4.** CpGPT performance by gastrointestinal cancer type.

| **Cancer type** | **Validation AUROC<sup>1</sup>** | **Test AUROC<sup>1</sup>** | **Test AUPRC<sup>1</sup>** |
| --- | --- | --- | --- |
| All | 0.9487<br>(0.8887–1.0000) | 0.9904<br>(0.9715–1.0000) | 0.9960<br>(0.9950–0.9970) |
| PDAC | 0.9069<br>(0.8255–0.9883) | 0.9600<br>(0.9383–0.9817) | 0.9792<br>(0.9658–0.9926) |
| HCC | 0.9667<br>(0.9134–1.0000) | 0.9792<br>(0.9458–1.0000) | 0.9917<br>(0.9783–1.0000) |
| ESCC | 0.9601<br>(0.9281–0.9921) | 1.0000<br>(1.0000–1.0000) | 1.0000<br>(1.0000–1.0000) |
| CRC | 1.0000<br>(1.0000–1.0000) | 1.0000<br>(1.0000–1.0000) | 1.0000<br>(1.0000–1.0000) |
| GC | 0.9556<br>(0.8844–1.0000) | 0.9833<br>(0.9566–1.0000) | 0.9833<br>(0.9566–1.0000) |
| EAC | 0.9000<br>(0.7868–1.0000) | 1.0000<br>(1.0000–1.0000) | 1.0000<br>(1.0000–1.0000) |

<sup>1</sup> Mean and 95% confidence interval (CI) are summarized across three split-runs. The 95% CIs were bounded to the valid metric range of 0 to 1.

Mean test AUROC was 0.9600 for pancreatic adenocarcinoma (95% CI, 0.9383–0.9817), 0.9792 for hepatocellular carcinoma (95% CI, 0.9458–1.0000), and 0.9833 for gastric cancer (95% CI, 0.9566–1.0000). Mean test AUROC was 1.0000 for esophageal squamous cell carcinoma, colorectal cancer, and esophageal adenocarcinoma, with 95% CIs of 1.0000–1.0000 across the three split-runs.

Mean test AUPRC was 0.9792 for pancreatic adenocarcinoma (95% CI, 0.9658–0.9926), 0.9917 for hepatocellular carcinoma (95% CI, 0.9783–1.0000), and 0.9833 for gastric cancer (95% CI, 0.9566–1.0000). Mean test AUPRC was 1.0000 for esophageal squamous cell carcinoma, colorectal cancer, and esophageal adenocarcinoma.

Although high cancer type-specific discrimination was observed, several cancer types had limited sample sizes, and the corresponding results reflect internal split-run performance rather than validation in independent cancer type-specific cohorts.

## Discussion

In this study, we developed and evaluated a DMR-informed framework for adapting CpGPT, a pretrained DNA methylation foundation model, to cfDNA-based gastrointestinal cancer classification. The technical sensitivity analysis using the previously published 896-CpG panGI panel provided information about the consistency of CpGPT fine-tuning across neighboring optimization settings. Under the base configuration, internal test AUROCs ranged from 0.9597 to 0.9927 across four fixed random seeds, while neighboring configurations achieved mean internal test AUROCs between 0.9661 and 0.9716. These results suggest that modest changes in learning rate, dropout, and condition-loss weight did not substantially alter performance within this dataset. However, because the analysis used a previously developed feature panel and samples from the same source cohort, it should not be interpreted as independent validation of either the panel or the resulting classifier.

A central feature of the framework was the integration of tumor tissue and plasma cfDNA methylation evidence before model fine-tuning. Tissue-derived DMRs identify epigenetic alterations associated with malignant transformation, but not all such alterations are readily detectable in plasma. Conversely, cfDNA-derived DMRs represent circulating methylation differences but may include signals arising from technical variation or non-tumor biological processes. Requiring support from both tissue and cfDNA analyses therefore provided a biologically constrained strategy for reducing the initial methylation feature space. This approach is conceptually consistent with the tissue-to-plasma biomarker discovery strategy used in EpiPanGI Dx, while extending that strategy to the adaptation of a pretrained methylation foundation model^[3]^.

The DMR-informed procedure reduced the candidate feature space to 20,499 CpG sites supported by both tumor-normal tissue differences and cancer-versus-control plasma differences. This feature set remained substantially larger than a clinically deployable targeted panel, but it provided a more focused input space than unrestricted genome-wide methylation profiling. The present analysis therefore demonstrates biologically informed candidate feature construction rather than selection of a finalized diagnostic panel. Additional feature attribution, stability-selection, redundancy-reduction, and experimental validation analyses will be required to determine whether a smaller subset of CpG sites can preserve predictive performance.

CpGPT achieved the highest mean test AUROC among the evaluated models. The strongest classical comparators were SVM radial and elastic net, whereas GBM and Random Forest produced lower mean AUROCs. These results suggest that both regularized linear and nonlinear kernel methods can extract substantial cancer-associated signal from the DMR-informed CpG set. The higher mean performance observed for CpGPT may reflect the benefit of representations learned during large-scale methylation pretraining and the model’s incorporation of methylation state, genomic position, and local sequence context^[4]^. Similar work with MethylGPT has also shown that transformer-based pretraining can generate reusable representations for downstream methylation-prediction tasks^[5]^.

Nevertheless, the model comparison should be interpreted descriptively. The confidence intervals summarized variability across three data split-runs rather than uncertainty estimated from participant-level resampling, and no formal paired hypothesis test was conducted to compare the models. The overlapping confidence intervals for several models also preclude concluding that CpGPT was statistically superior based on the present analysis alone. Larger evaluation cohorts and repeated or nested resampling designs would permit more precise estimation of between-model performance differences.

The panGI technical sensitivity analysis provided complementary evidence that CpGPT performance was not confined to a single data split or exact hyperparameter setting. Under the base configuration, test AUROCs ranged from 0.9597 to 0.9927 across four fixed random seeds, and neighboring configurations retained mean test AUROCs between 0.9661 and 0.9716. The relatively narrow range of high performance across learning-rate, dropout, and condition-loss settings supports the technical stability of fine-tuning in this dataset. This finding is relevant to applications involving targeted methylation panels, in which modest changes in training conditions should not result in substantial loss of discrimination.

Cancer type-specific analyses showed high mean test AUROC for each of the six gastrointestinal cancer types. Mean AUROC was 0.9600 for pancreatic adenocarcinoma, 0.9792 for hepatocellular carcinoma, and 0.9833 for gastric cancer. Esophageal squamous cell carcinoma, colorectal cancer, and esophageal adenocarcinoma each had a mean test AUROC of 1.0000 across the evaluated split-runs. These findings suggest that the aggregate cancer-versus-control performance was not driven exclusively by one cancer type. However, the perfect estimates should not be interpreted as evidence of perfect population-level discrimination. Several cancer types had small total sample counts, and individual test partitions contained only a subset of those samples. Consequently, a small number of correctly ranked observations could produce an AUROC of 1.0000.

The observed performance is broadly consistent with prior evidence that cfDNA methylation can distinguish gastrointestinal cancers from non-cancer controls^[3]^ and with multicancer studies demonstrating that methylation signals can support both cancer detection and tissue-of-origin inference^[2]^. Direct numerical comparison with previous studies is difficult because cohort composition, control selection, methylation assays, feature panels, data partitions, and classification endpoints differ. The current study should therefore be viewed as a computational proof of concept rather than a direct comparative evaluation against previously published clinical assays.

The results also illustrate a potential role for foundation models in settings where labeled cfDNA cohorts are modest in size. Rather than learning all methylation representations de novo from several hundred samples, CpGPT begins with representations learned from large-scale methylation pretraining^[4]^. Such transfer may improve data efficiency and support the integration of positional and sequence-context information that is not explicitly modeled by standard feature-vector classifiers. However, the present study did not directly isolate the contribution of pretraining, architecture, or sequence context. Ablation studies comparing the pretrained model with randomly initialized or reduced-context variants would be needed to determine which model components account for the observed performance.

Several limitations should be considered. First, this was a retrospective analysis of a single publicly available cfDNA dataset. Although performance was examined across multiple split-runs, all training and evaluation samples originated from the same source cohort and assay workflow. The study therefore lacks independent external validation across institutions, populations, sample-processing protocols, sequencing batches, and laboratory platforms.

Second, the cohort contained only 46 non-cancer controls. The precision of specificity-related performance estimates is consequently limited, and the controls may not represent the full clinical spectrum in which the model would be applied. Future evaluations should include larger numbers of individuals without cancer, including participants with benign gastrointestinal disease, inflammatory conditions, liver disease, premalignant lesions, and other cancers that could generate overlapping cfDNA methylation signals.

Third, the present analyses focused primarily on discrimination. AUROC and AUPRC do not establish that predicted probabilities are well calibrated or that a model provides clinical benefit at relevant decision thresholds. Future studies should assess calibration, sensitivity at prespecified specificity levels, predictive values under realistic disease prevalence, decision-curve performance, and analytical reproducibility. Transparent reporting of evaluation datasets, subgroup performance, calibration, and external validation is also consistent with current TRIPOD+AI recommendations for prediction-model studies^[16]^.

Fourth, the effective test sample sizes were small, particularly in the cancer type-specific analyses. Because each approximately 10% test partition contained only a small number of non-cancer controls and, for some cancer types, only one or a few cancer cases, an AUROC of 1.000 could result from the correct ranking of a very limited number of observations. Cancer type-specific results should therefore be regarded as exploratory and hypothesis-generating rather than precise estimates of population-level discrimination. The present study was not powered to evaluate performance by cancer stage, tumor burden, tumor fraction, age, sex, or other clinically relevant characteristics.

Fifth, the biological interpretability of the candidate set was inferred from the DMR-integration procedure rather than demonstrated through model-specific attribution analyses. The overlap of tissue and plasma signals increases biological plausibility, but it does not establish which CpGs drove individual predictions or whether the model learned cancer-specific regulatory mechanisms. Genomic annotation, pathway enrichment, attention or attribution analysis, feature-stability assessment, and experimental confirmation will be needed to characterize the biological relevance of the CpGs used by the model.

Finally, the reported uncertainty intervals were derived from variation across only three or four data-split or random-seed runs. Because the same participants were reused across runs, these runs do not represent independent participant-level samples. The intervals, therefore, summarize run-to-run variability rather than conventional sampling uncertainty and should not be interpreted as precise confidence bounds for clinical deployment performance. This limitation is particularly important for metrics approaching 1.000, for which bounded intervals may appear artificially narrow. Future studies should estimate uncertainty using participant-level bootstrap procedures or other prespecified resampling approaches in independent evaluation cohorts.

In conclusion, DMR-informed fine-tuning of CpGPT achieved high internal discrimination for cfDNA-based gastrointestinal cancer classification and the highest mean test AUROC among the evaluated transformer and classical machine learning approaches. Integrating tissue-derived and cfDNA-derived methylation signals provided a biologically constrained candidate feature set for foundation-model adaptation. These findings support further investigation of methylation foundation models for liquid-biopsy applications, while underscoring the need for external validation, clinically representative controls, calibration assessment, model-interpretation analyses, and reduction of the candidate feature set before targeted assay development.

## Data Availability

N/A

## Notes

### Competing Interest Statement

The authors have declared no competing interest.

